# Real-world retention of newer antiseizure medications in children with epilepsy – a nationwide study

**DOI:** 10.64898/2026.01.15.26344172

**Authors:** André Idegård, Ronny Wickström, Johan Zelano

**Author notes:** equal contribution (Ronny Wickström, Johan Zelano).

## Abstract

**Background:** Children with epilepsy are rarely included in clinical drug trials, delaying approval and introduction of new antiseizure medications (ASMs). To circumvent this we used population-wide register data to study retention, an intergrated meassure of effect and tolerability, of newer ASMs used in children in Sweden; brivaracetam, cenobamate, lacosamide, and perampanel.

**Methods:** All cases of incident epilepsy and subsequent ASM treatment 2007-2021 were identified using national registers. Retention rates of newer ASMs were calculated using Kaplan-Meier analyses. Cox regression was used to investigate how factors associated with therapy-resistant epilepsy affected risk of discontinuing each drug and to compare the risk of discontinuing each of the newer ASMs to levetiracetam, the most common ASM, in patients matched for age at epilepsy onset and number of previously tried ASMs.

**Results:** Out of 16431 included 54% were male, median age 7 years [Q1-Q3:3-12]. Developmental or intellectual disorders, followed by neurological malformations, were the most common comorbidities. In 2023 ∼10% of all ongoing treatments started <18years were brivaracetam, cenobamate, lacosamide, or perampanel. Brivaracetam, cenobamate, and lacosamide were all used off-label before authorization in children. Brivaracetam had the highest three-year retention of all antiseizure medications, 67%(95%CI:60-76) for the whole cohort, lacosamide and lamotrigine the highest five-year retentions: 40%(95%CI:36-45) and 45%(95%CI:43-46). perampanel had the lowest four-year retention. Retentions in the youngest were modest. In those 1-5years brivaracetam had among the highest three-year retention: 64%(95%CI:52-79), followed by lamotrigine and valproate. Lacosamide and lamotrigine had the highest five-year retentions: 40%(95%CI:33-49) and 42%(95%CI:39-44). Brivaracetam had the highest of all three-year retentions in those 6-11: 74%(95%CI:63-87) and lacosamide among the highest five-year retentions: 48%(95%CI:40-57), together with lamotrigine. In the eldest brivaracetam and lamotrigine had the highest retentions: 68%(95%CI:51-91) and 62%(95%CI:60-64), lacosamide had lower five-year retention than levetiracetam and lamotrigine. In Cox regression users of newer antiseizure medications, age-matched to levetiracetam-users with equal number previous antiseizure medications, the hazard ratio for discontinuing brivaracetam was 0.5(95%CI:0.4-0.8), perampanel 2.0(95%CI:1.2-3.2), and lacosamide 1.0(95%CI:0.8-1.2).

**Conclusion:** Brivaracetam show higher or similar retention and lacosamide similar retention as levetiracetam and lamotrigine, suggesting these could be treatment options in early epilepsy. Off-label use is common stressing the need for pediatric clinical trials.

## Introduction

The development of new antiseizure medications (ASMs) has accelerated in recent decades, driven by advances in molecular pharmacology and drug design. This offers more therapeutic options although the time from drug development is still long due to regulatory and safety requirements. Newer ASMs are typically reserved for severe drug-resistant epilepsy, in which the risk-benefit ratio favors testing new drugs despite a relative scarcity of safety data.^1^

Pediatric populations are rarely included in clinical trials, limiting the availability of efficacy and safety data, including long-term safety, and thereby approval for pediatric use. To address this problem, the European Medicines Agency (EMA) now requires all applications for marketing authorization for new medicines to include a pediatric investigation plan aimed at ensuring that the necessary data are obtained through studies in children.^2^ Still, the authorization for newer drugs including ASMs is substantially slower in children.^3^

For the regulatory and safety reasons described above, newer ASMs are only very slowly introduced as first or second therapy in adults.^4^ The situation is likely to be worse in children, although off-label prescription seems common.^3^ Tracking ASM use through register data allows for nation-wide study of newer ASMs, including estimation of early use as well as retention rates, an integrated measurement om efficacy and tolerability.^5^ Using comprehensive national register data of all incident pediatric epilepsy patients in Sweden 2007-2023 we aimed to compare newer ASMs to older. We focused our analysis on brivaracetam (BRV), cenobamate (CNB), lacosamide (LCM), and perampanel (PER) and compared them to common older ASMs. Our hypothesis, based on findings in adults, was that the newer ASMs would have at least similar retention as more established ASMs.^4^

## Methods

### Registers and definitions

The National Patient Register contains all in- and outpatient hospital diagnoses in Sweden. Reporting to the patient register is mandatory and is done by all state-funded care-providers, which make up the vast majority of Swedish healthcare. However some privately funded care providers do not report to the registers, the exact proportion is unknown but constitute a small proportion of the Swedish healthcare system.^6^ Overall the quality of the patient register is high and most diagnoses have high positive predictive values.^7^

The Cause of Death Register contains all deaths in Sweden. The Register receives data from the Swedish Tax Agency with little to no missingness. In an estimated 2-3% of cases the causes of death are missing or of poor quality.^8^

The National Prescribed Drugs Register contains all prescribed medications dispensed from all pharmacies in Sweden since July 1, 2005. The reporting is automated and connected to the system that reimburses the pharmacies for the subsidized part of medication costs, it is therefore in the pharmacies own interest to report and the coverage is estimated to 100%.^9^ Prescribed medications are subsidized by the government. The maximum patient-cost has historically been low and was 2900 SEK (310 USD/265 EUR) in 2024, and fully subsidized in children with no patient-cost.^10^ The cost for pharmacies if not reporting to the register would thus be immense.

Epilepsy was defined as having an International Classification of Disease 10 (ICD-10) code G40 and at least one dispensation of an ASM, defined as Anatomical Therapeutical Chemical code N03A. Epilepsy onset was defined as the first occurrence of any seizure-related code; G40, G41, or R568. Definitions of comorbidities are available in supplementary eTable 1. Information on EMA approval of ASMs was collected from Electronic Medicines Compendium (www.medicines.org.uk).

### Cohort

Through cross-linking the national registers, all children aged one month to 17 years in Sweden with epilepsy and subsequent ASM treatment (n = 16,435) 2007-2021 were identified. As our data spans through 2005-2023, this allowed analysis two years prior to any seizure or ASM-treatment to ensure incident epilepsy, and a minimum two years of follow-up for all individuals included. Three patients with suspected reused personal numbers and one with inconsistent data (dead before inclusion) were excluded.

### Statistical analyses

A previously described algorithm was used to track ASM usage in the register data.^11^ Briefly, a patient was considered on treatment if they had a minimum of two dispensations of the drug within 365 days. Discontinuation was defined as more than 365 days between dispensations. The last day of treatment was then set as 90 days after the last dispensation, the time one dispensation normally lasts.

Retention rates were estimated using Kaplan-Meier analysis. Time was calculated as time from start of the ASM to discontinuation (event), death, or last day of the study (December 31, 2023). Kaplan-Meier analyses were censored if fewer than ten individuals remained at risk. The ASM had to be started before age 18 to be included in the analyses.

Since the newer ASMs were often used after a greater number of previous ASMs than the older ones, we also performed matched analyses. The patients using the new ASM were matched to patients using Levetiracetam (LEV) after the same number of previous ASMs and who had never used the newer ASM, with matching for age at epilepsy onset. Cox proportional hazards modelling, stratified by number of previous ASMs, was conducted individually for each of BRV, LCM, and PER with LEV as a reference in these matched cohorts.

Cox Proportional Hazards modelling was also used to evaluate how known predictors of therapy resistant epilepsy; age, sex, brain infections and tumors, inborn errors of metabolism, congenital neurological malformations, developmental and intellectual disabilities, and status epilepticus; influenced discontinuation.^12^ The analyses were done separately for BRV, LCM, and PER since the same individual might have used multiple of them, thereby violating the model assumptions if analyzed together.

For all incomplete dates for hospital visits in the patient register the year in question was available, the date was then defined as first of July the corresponding year. Regarding birth dates, where we only had access to year and month, the first day of the month was set as date of birth. Incomplete death dates were set as the last of the month if available, or else at end of the year. No other imputations were made.

All analyses were conducted using R version 4.5.1.^13^ Kaplan-Meier analysis and Cox proportional hazards modelling were conducted using the *survival* package, the proportional hazards assumptions were controlled using the *coxzph* function.^14^ Matching of LEV-users to those of the newer ASMs were done using the *MatchIt* package.^15^

### Ethics

The study was approved by the Swedish Ethical Review Authority, approval number 2023-05598-01.

### Data availability

The underlying register data cannot be shared by the authors due to Swedish privacy regulations but are available from the register holders following application.

## Results

There were 16,431 patients with pediatric-onset epilepsy after 1 January 2007, 54% were male (Table 1). Baseline characteristics were described for age strata. Congenital neurological malformations or inborn errors of metabolism were more commonly seen in children with epilepsy onset before one year of age, whereas intellectual or developmental disabilities were more commonly seen in children with epilepsy-onset before age six.

**Table 1.** Study cohort.

| **n (%)** | **All** | **1mo-<1yr** | **1-5yrs** | **6-11yrs** | **>12yrs** |
| --- | --- | --- | --- | --- | --- |
| **n** | 16431 | 1900 | 4834 | 5275 | 4422 |
| **Time in study (years), median [Q1-Q3]** | 9 [5-12] | 9 [5-12] | 9 [5-12] | 8 [5-12] | 9 [5-13] |
| **Age, median [Q1-Q3]** | 7 [3-12] | 0 [0-0] | 3 [2-4] | 8 [7-10] | 15 [13-16] |
| **Male sex** | 8833 (54) | 972 (51) | 2702 (56) | 2889 (55) | 2270 (51) |
| **Female sex** | 7598 (46) | 928 (49) | 2132 (44) | 2386 (45) | 2152 (49) |
| **Status epilepticus** | 364 (2) | 61 (3) | 178 (4) | 80 (2) | 45 (1) |
| **Brain infection** | 332 (2) | 45 (2) | 109 (2) | 85 (2) | 93 (2) |
| **Brain tumor** | 154 (1) | 13 (1) | 25 (1) | 45 (1) | 71 (2) |
| **Developmental or intellectual disorder** | 5057 (31) | 763 (40) | 1897 (39) | 1419 (27) | 978 (22) |
| **Inborn errors of metabolism** | 313 (2) | 84 (4) | 110 (2) | 79 (1) | 40 (1) |
| **Congenital neurological malformation** | 1020 (6) | 332 (17) | 387 (8) | 186 (4) | 115 (3) |
| **Tuberous sclerosis** | 135 (1) | 64 (3) | 50 (1) | 13 (0) | 8 (0) |

We first investigated how newer ASMs were prescribed during the study period. They were rarely used as first or second ASMs but use of BRV and LCM as second therapy increased slightly towards the end of the study period (Figure 1A-B). For the majority of patients, the newer drugs were started only after having tried multiple other ASMs, and often as the patients were older (eFigure 1). Overall, the use of the newer ASMs that were started before 18 years of age increased throughout the study period to about 10% of all ongoing therapies in 2023 (Figure 1C). Figure 1D lists approval dates for the newer ASMs, including authorization for pediatric use. BRV, CNB, and LCM were all used in the cohort before authorization for pediatric use.

**Figure 1.**
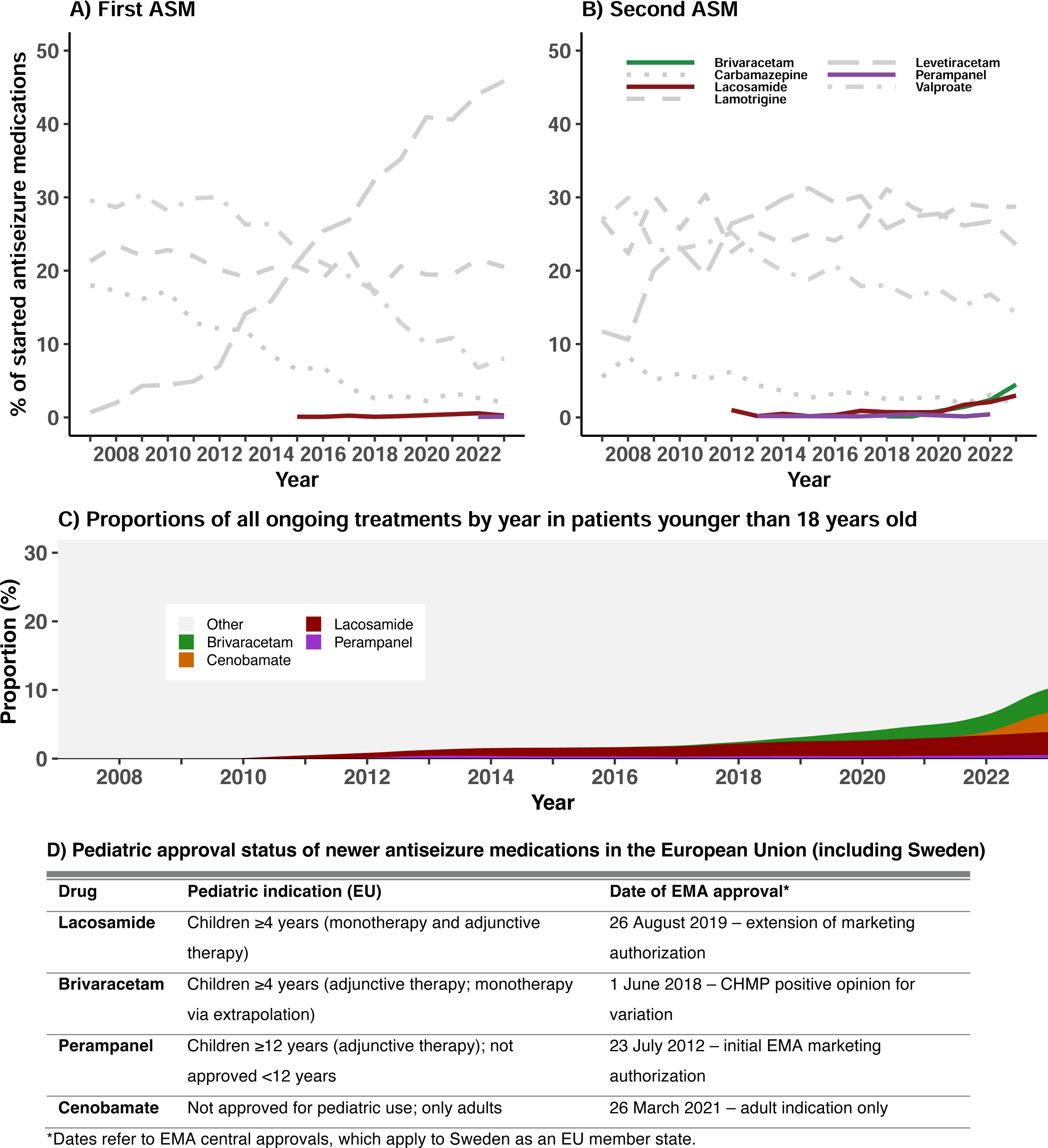
Prescription patterns for first (A) and second (B) antiseizure medication (ASM), proportions of all ongoing treatments, started before 18 years of age, by year (C), and pediatric approval dates of the newer antiseizure medications (D).

Retention rates of ASMs were studied for the first episode the newer drugs were used in each patient (Figure 2, Table 2). Of the newer drugs only LCM had enough users to calculate five-year retention rates. Overall, BRV had the highest retention after three years (67%, 95% CI: 60-76), followed by LTG (58%, 95% CI: 57-59). LCM and LTG had the highest retention at five years at 40% (95% CI: 36-45) and 45% (95% CI: 43-46) respectively. PER had the lowest retention rate after four years (20%, 95% CI: 14-31). Unfortunately too few children had used CNB to calculate retention.

**Figure 2.**
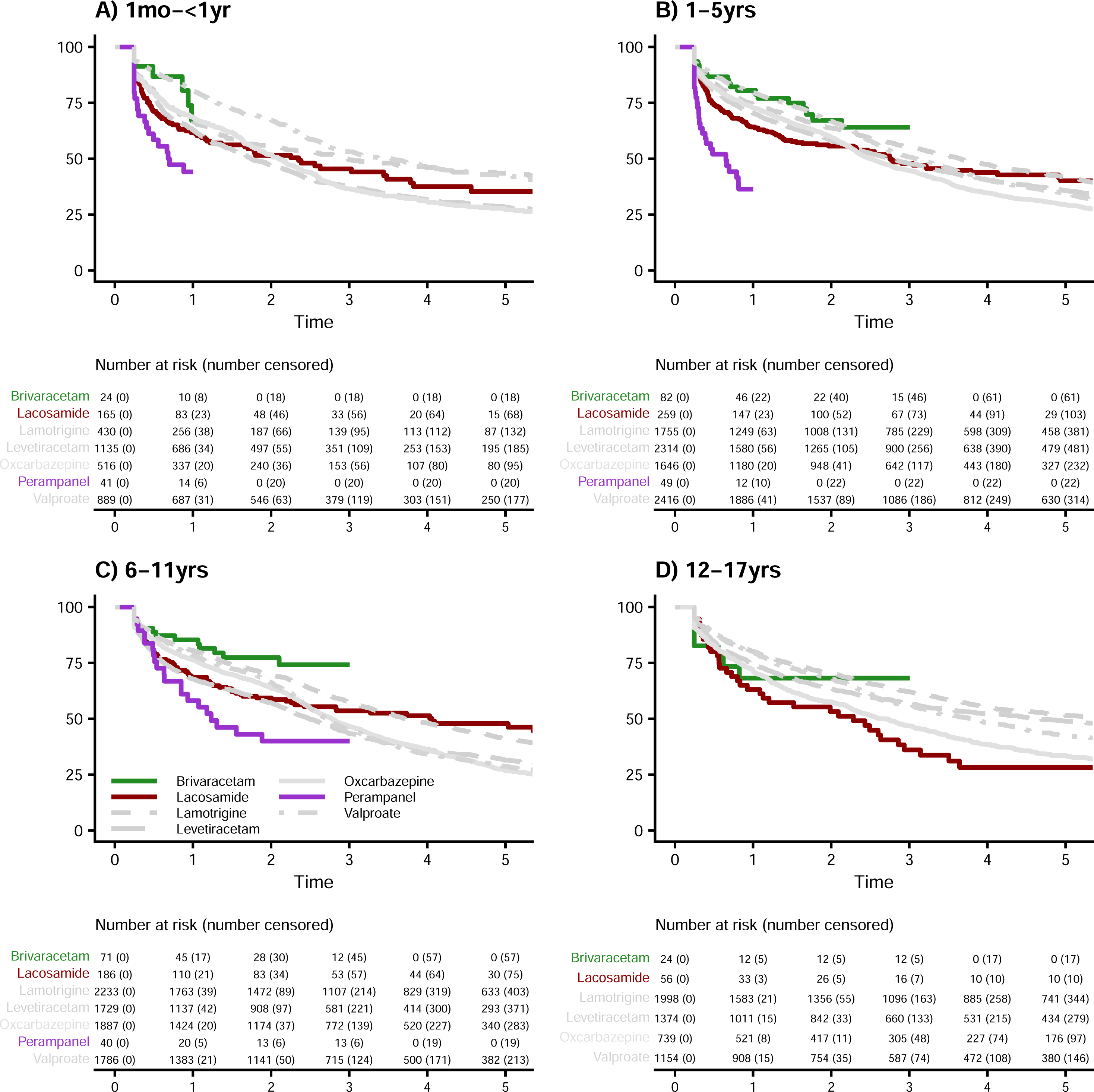
Kaplan-Meier curves of retention rates of ASMs the first time they were used in each patient.

**Table 2.** Retention rates for all antuseizure medications started before age 18, sorted by retention rate at one year.

| All | 1 year | 2 years | 3 years | 4 years | 5 years |
| --- | --- | --- | --- | --- | --- |
| Valproate | 79.4 (78.4-80.4) | 66.9 (65.7-68.1) | 50.5 (49.2-51.8) | 40.9 (39.7-42.2) | 35.3 (34.1-36.6) |
| Brivaracetam | 79.2 (73.4-85.6) | 69.7 (62.7-77.6) | 67.3 (59.9-75.7) |  |  |
| Lamotrigine | 77.9 (76.8-78.9) | 67.3 (66.1-68.5) | 57.9 (56.6-59.1) | 50.0 (48.7-51.3) | 44.7 (43.4-46.0) |
| Oxcarbazepine | 73.5 (72.3-74.8) | 60.1 (58.8-61.6) | 44.9 (43.5-46.4) | 35.4 (34.0-36.9) | 28.8 (27.4-30.2) |
| Levetiracetam | 69.3 (68.2-70.4) | 57.2 (56.0-58.4) | 46.9 (45.7-48.2) | 40.5 (39.2-41.8) | 36.3 (35.0-37.6) |
| Lacosamide | 64.7 (61.1-68.6) | 55.5 (51.6-59.6) | 47.6 (43.5-52.0) | 42.8 (38.5-47.5) | 40.0 (35.6-44.9) |
| Perampanel | 43.6 (35.7-53.3) | 30.1 (22.7-39.9) | 26.6 (19.4-36.4) | 20.3 (13.5-30.6) |  |
| 1m-<1yr | 1 year | 2 years | 3 years | 4 years | 5 years |
| Valproate | 80.4 (77.8-83.1) | 67.3 (64.3-70.6) | 53.0 (49.7-56.6) | 46.6 (43.2-50.2) | 42.3 (38.9-46.0) |
| Oxcarbazepine | 68.7 (64.7-72.8) | 51.8 (47.5-56.4) | 36.5 (32.4-41.2) | 30.8 (26.8-35.4) | 27.1 (23.2-31.7) |
| Brivaracetam | 67.1 (48.1-93.7) |  |  |  |  |
| Lamotrigine | 66.9 (62.5-71.6) | 55.7 (51.0-60.8) | 49.5 (44.6-54.9) | 46.1 (41.1-51.6) | 43.5 (38.5-49.2) |
| Levetiracetam | 62.8 (60.0-65.7) | 47.2 (44.4-50.3) | 38.0 (35.2-41.0) | 31.8 (29.0-34.8) | 28.3 (25.6-31.3) |
| Lacosamide | 61.7 (54.4-69.9) | 51.5 (43.8-60.6) | 45.4 (37.4-55.3) | 37.5 (29.0-48.6) | 35.3 (26.6-47.0) |
| Perampanel | 44.1 (30.6-63.5) |  |  |  |  |
| 1-5yrs | 1 year | 2 years | 3 years | 4 years | 5 years |
| Brivaracetam | 80.6 (71.9-90.3) | 67.1 (55.6-81.0) | 64.1 (52.0-79.0) |  |  |
| Valproate | 79.6 (78.0-81.2) | 66.7 (64.9-68.7) | 50.8 (48.8-52.9) | 40.5 (38.5-42.6) | 34.5 (32.5-36.6) |
| Lamotrigine | 74.3 (72.2-76.4) | 63.7 (61.5-66.1) | 55.4 (53.0-57.9) | 47.4 (45.0-50.0) | 41.7 (39.2-44.3) |
| Oxcarbazepine | 72.7 (70.6-74.9) | 59.6 (57.3-62.1) | 44.7 (42.3-47.2) | 34.7 (32.4-37.2) | 29.4 (27.1-31.8) |
| Levetiracetam | 70.4 (68.5-72.3) | 58.3 (56.3-60.4) | 47.9 (45.8-50.0) | 40.4 (38.4-42.6) | 35.8 (33.7-38.0) |
| Lacosamide | 64.2 (58.5-70.5) | 55.6 (49.6-62.4) | 47.8 (41.5-55.2) | 43.8 (37.2-51.5) | 40.1 (33.1-48.6) |
| Perampanel | 36.4 (24.3-54.6) |  |  |  |  |
| 6-11yrs | 1 year | 2 years | 3 years | 4 years | 5 years |
| Brivaracetam | 85.3 (76.8-94.7) | 77.4 (67.1-89.1) | 74.1 (62.9-87.4) |  |  |
| Lamotrigine | 80.6 (78.9-82.2) | 69.4 (67.5-71.4) | 57.5 (55.4-59.6) | 48.1 (46.0-50.3) | 41.3 (39.1-43.5) |
| Valproate | 78.5 (76.6-80.4) | 66.2 (64.1-68.5) | 45.1 (42.8-47.6) | 34.2 (31.9-36.5) | 28.8 (26.6-31.1) |
| Oxcarbazepine | 76.4 (74.5-78.3) | 63.8 (61.7-66.0) | 46.7 (44.4-49.0) | 36.0 (33.8-38.4) | 26.9 (24.8-29.2) |
| Lacosamide | 68.6 (62.0-75.9) | 59.3 (52.3-67.3) | 53.6 (46.2-62.1) | 51.3 (43.8-60.2) | 47.8 (40.0-57.1) |
| Levetiracetam | 67.8 (65.6-70.0) | 57.2 (54.9-59.6) | 43.3 (41.0-45.9) | 36.3 (33.9-38.8) | 31.5 (29.1-34.0) |
| Perampanel | 58.1 (43.9-76.9) | 40.0 (26.5-60.3) | 40.0 (26.5-60.3) |  |  |
| 12-17yrs | 1 year | 2 years | 3 years | 4 years | 5 years |
| Lamotrigine | 80.2 (78.4-82.0) | 70.3 (68.3-72.4) | 62.1 (60.0-64.3) | 55.2 (53.0-57.5) | 51.4 (49.2-53.8) |
| Valproate | 79.9 (77.6-82.2) | 68.0 (65.3-70.8) | 56.2 (53.4-59.2) | 48.2 (45.3-51.3) | 42.4 (39.6-45.5) |
| Levetiracetam | 74.5 (72.3-76.9) | 63.3 (60.8-65.9) | 56.7 (54.1-59.4) | 52.4 (49.8-55.2) | 48.9 (46.2-51.8) |
| Oxcarbazepine | 71.5 (68.3-74.8) | 57.6 (54.1-61.3) | 46.8 (43.2-50.6) | 38.5 (35.1-42.3) | 33.5 (30.0-37.3) |
| Brivaracetam | 68.2 (51.1-90.9) | 68.2 (51.1-90.9) | 68.2 (51.1-90.9) |  |  |
| Lacosamide | 63.1 (51.4-77.4) | 53.2 (41.3-68.5) | 36.1 (24.9-52.3) | 28.2 (17.7-45.0) | 28.2 (17.7-45.0) |

Relatively few patients with epilepsy onset before one year of age had used the newer ASMs, but LCM had lower retention than VPA or LTG in those who had. In children with epilepsy onset between one and five years BRV had among the highest retention rates after three years, 64% (95% CI: 52-79). LCM and LTG had the highest five-year retention in these patients: 40%, (95% CI: 33-49) and 42% (95% CI: 39-44). In those with epilepsy onset between six and eleven years, BRV had the highest three-year retention rate, 74% (95% CI: 63-87), higher than both LTG (58%, 95% CI: 55-60) and VPA. LCM and LTG had the highest five-year retentions in these patients, 48% (95% CI: 40-57) and 41% (95% CI: 39-44), respectively. In children aged 12 or older at epilepsy onset, BRV had among the higher three-year retentions (68%, 95% CI: 51-91). In sex-stratified analyses of those over 11 years at epilepsy onset, female patients generally had lower retention rates than males. LCM had similar two-year retention as other ASMs in males but relatively low retention in females (eTable 2).

To compensate for the differences in number of previous ASMs between the newer and older ASMs, we also performed Cox proportional hazards modelling with LEV as reference in which users of newer ASMs were matched for age at epilepsy onset and number of previous ASMs. The risk of discontinuing BRV was lower (HR 0.5, 95% CI: 0.4-0.8) compared to LEV whereas the risk of discontinuing PER was higher (HR 2, 95% CI: 1.2-3.2, Figure 3). The risk of discontinuing LCM was similar to that of LEV.

**Figure 3.**
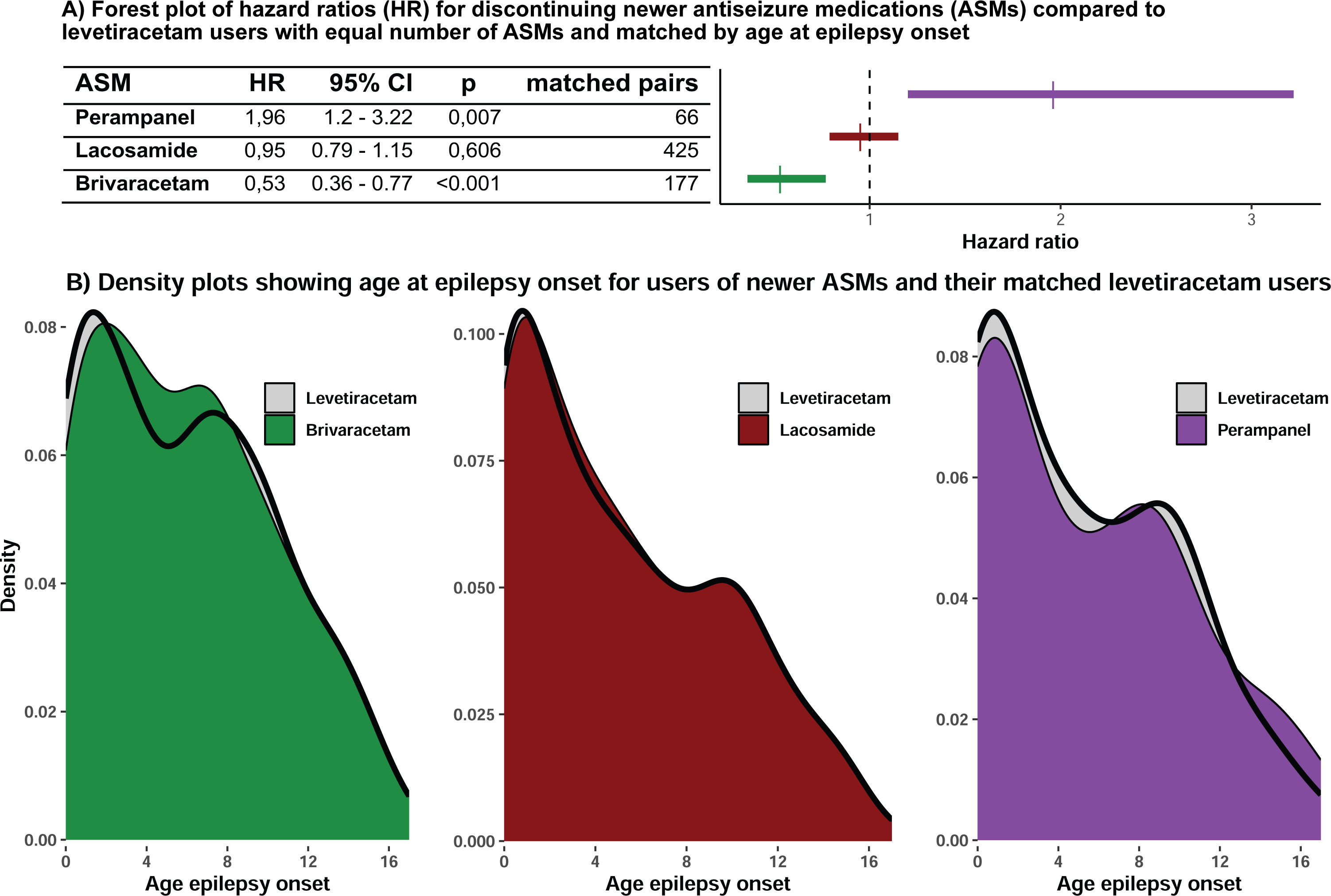
Hazard ratios for discontinuing newer antiseizure medications (ASMs) compared to levetiracetam in patients with the same number of previous ASMs, matched by age at epilepsy onset (A) and density plots showing overlap of age at epilepsy onset between the users of the newer ASMs and their matched levetiracetam controls (B).

In Cox proportional hazards modelling, assessing whether patient characteristics associated with therapy resistant epilepsy affected the risk of discontinuing the ASMs, intellectual or developmental disorders were associated with increased risk of discontinuing LCM and brain infections with discontinuation of PER in unadjusted models, although confidence intervals were wide (eTable 3).

## Discussion

In this nation-wide study of all patients with pediatric epilepsy and subsequent ASM-treatment in Sweden 2007-2021 we show that newer ASMs are increasingly used in children, although rarely as first or second medication. Similar to previous findings, off-label use was common.^3^ LCM was used nine years ahead of its approval in children, and CNB has been used since 2021 despite still not being approved for pediatric use. BRV had high retention rates, similar to, or higher than, common ASMs such as LEV, LTG and VPA.

Our findings are well in line with the existing literature. Recent randomized trials have also reported that LCM has a favorable safety profile compared to oxcarbazepine and PER, in line with the relatively high long-term retention rates we find.^16,17^ Previously reported one-year retention rates of LCM in clinical studies range from 40-90%.^18–21^ In our cohort 65% of lacosamide users were still using the drug after one year. Previously reported retention rates of BRV are generally lower than our findings; an open-label follow-up trial reported a four-year retention rate of 58%,^22^ compared to 67% in our cohort and a meta-analysis including 9 studies reported one-year retention of 66% compared to the 79% in our cohort.^23^

The PER retention rates in our cohort are surprisingly low, in particular in younger children, compared to systematic reviews of mainly Asian studies,^24,25^ although one British multicenter study found similarly low retention rates.^26^ The tendency of PER users having tried multiple other ASMs prior to treatment might explain part of this difference, also seen in other studies.^27^.

The overall differences between our findings and those previously reported might stem from differences in study design. While we include everyone with epilepsy, both therapy-resistant and not, and regardless of epilepsy-type, previous studies have mostly covered therapy refractory epilepsy and often specific types of epilepsy. Considering that the majority of our cohort used newer ASMs after having tried multiple others, and the high proportions of intellectual or developmental disorders, many patients likely fulfill the criteria for drug-resistant epilepsy.^12,28^ As the registers do not contain information on why medications were discontinued the exact proportion meeting the criteria is however unknown.

When comparing the retention rates of the different ASMs within the study it is important to consider that the older ASMs were often used as one of the first ASMs while the newer ones were often used after having tried multiple others. This likely affects the reasons for discontinuation and therefor retention rates; patients who have tried multiple others might be more willing to accept some side effects and to try an ASM for longer period before deciding whether it is effective or not. The Cox models where BRV-, LCM-, and PER-users were matched to LEV-users with the same number of previous ASMs do however suggest that our findings are consistent despite this potential limitation.

The findings of the present study add to previous reports by the comprehensive and unbiased inclusion of all patients in Sweden, the major strength of register studies. This also make the results more generalizable than many clinical trials that focuses on specific patient groups as opposed to including every child with epilepsy. As prescription drugs for children in Sweden are fully subsidized by the government, with no patient-cost, there is likely no bias in ASM-choice based on medication costs. However, there are also limitations in register-based method. The temporal resolution is relatively poor, and we are unable to separate withdrawals for lack of efficacy from adverse events. On the other hand, retention is a recommended outcome for clinical ASM trials.^5^ Another possible limitation is misclassification, although the combination of a diagnostic code for epilepsy and ASM prescriptions is the most specific for cases of epilepsy and the validity of ICD-10 codes, including G40 for epilepsy, in Swedish registers is generally high^7,29,30^. We are also unable to measure adherence, i.e., whether patients have actually taken the drugs prescribed.

Despite these shortcomings, our study illustrates the potential for big data to collect all real-world experience on new ASMs from an entire country. If multiple registers from different countries with large registers and insurance databases are combined in multicenter efforts – big data could offer a way of accelerating dissemination of the newest ASMs to broad patient groups. Our findings also underline the need for randomized trials of newer ASMs in pediatric-onset epilepsy. If newer ASMs have as high or even higher retention than older ones, they could provide a valuable addition to the therapeutic arsenal in new-onset epilepsy.

## Conclusion

We conclude that BRV appear to have equal to or higher retention rates than, and that LCM has retention rates comparable to older ASMs as treatment for pediatric-onset epilepsy. Beyond this we demonstrate that off-label prescription is still common in children, once again illustrating the need to include children early on in pharmaceutical trials.

