## Supplemental material for "Real-world retention of newer antiseizure medications in children with epilepsy – a nationwide study"

eTable 1. International Classification of Disease version 10 (ICD-10) codes defining conditions and comorbidities.

| Diagnose | ICD-10 |
| --- | --- |
| Epilepsy | G40 |
| Seizure | G40, G41, R568 |
| Status epilepticus | G41 |
| Brain tumor | C71, C793, D430, D32, D330 |
| Inborn disorder | Q0, Q1, Q2, Q3, Q4, Q5, Q6, Q7, Q80, Q81, Q82, Q83,  Q84, Q850, Q858, Q859, Q86, Q87, Q88, Q89, Q9 |
| Intellectual or developmental disorder | F7, F84 |
| Tuberous sclerosis | Q851 |

eTable 2. Retention rates of those age 12 or older stratified by sex.

| Male | 1 year | 2 years | 3 years | 4 years | 5 years |
| --- | --- | --- | --- | --- | --- |
| Valproate | 84.5 (82.1-87.0) | 73.9 (71.0-76.9) | 61.6 (58.4-65.0) | 53.0 (49.6-56.6) | 47.6 (44.2-51.3) |
| Lamotrigine | 82.2 (79.6-85.0) | 72.6 (69.5-75.9) | 65.7 (62.4-69.2) | 59.4 (55.9-63.1) | 56.1 (52.5-59.9) |
| Oxcarbazepine | 75.4 (71.2-79.9) | 62.6 (57.9-67.7) | 52.3 (47.5-57.7) | 43.0 (38.2-48.5) | 38.4 (33.6-44.0) |
| Levetiracetam | 73.8 (70.3-77.5) | 65.6 (61.8-69.6) | 58.0 (54.0-62.3) | 54.2 (50.1-58.6) | 51.8 (47.7-56.4) |
| Lacosamide | 73.7 (58.7-92.5) | 65.7 (49.8-86.7) |  |  |  |
| Female | **1 year** | **2 years** | **3 years** | **4 years** | **5 years** |
| Lamotrigine | 78.9 (76.6-81.2) | 68.9 (66.3-71.5) | 59.9 (57.1-62.7) | 52.5 (49.7-55.5) | 48.6 (45.7-51.6) |
| Levetiracetam | 75.1 (72.1-78.1) | 61.7 (58.4-65.2) | 55.8 (52.4-59.4) | 51.2 (47.7-54.9) | 46.9 (43.4-50.7) |
| Oxcarbazepine | 67.2 (62.5-72.3) | 52.3 (47.3-57.7) | 40.9 (36.1-46.4) | 33.8 (29.1-39.2) | 28.2 (23.7-33.6) |
| Valproate | 66.9 (61.8-72.4) | 51.5 (46.2-57.5) | 41.2 (36.0-47.2) | 35.0 (29.9-40.8) | 28.3 (23.6-34.0) |
| Lacosamide | 52.9 (37.2-75.4) | 41.6 (26.6-65.0) |  |  |  |

eTable 3. Hazard ratios (HR) for discontinuing each antiseizure medication, calculated with Cox Proportional Hazards Modelling.

|  | Unadjusted | | | Adjusted | | |
| --- | --- | --- | --- | --- | --- | --- |
| BRV | **HR** | **95% CI** | **p** | **HR** | **95% CI** | **p** |
| Age epilepsy onset | 1.01 | 0.95 - 1.08 | 0,711 | 1.03 | 0.96 - 1.11 | 0,44 |
| Female | 0.85 | 0.49 - 1.47 | 0,563 | 0.83 | 0.47 - 1.45 | 0,506 |
| Nr previous ASMs | 1.04 | 0.97 - 1.13 | 0,273 | 1.07 | 0.98 - 1.17 | 0,154 |
| Brain infection | 1.75 | 0.55 - 5.65 | 0,346 | 2.08 | 0.44 - 9.77 | 0,355 |
| Intellectual or developmental disorder | 0.96 | 0.56 - 1.65 | 0,879 | 0.8 | 0.42 - 1.51 | 0,486 |
| Neurological malformation | 0.66 | 0.24 - 1.84 | 0,427 | 0.68 | 0.23 - 1.96 | 0,473 |
| Status epilepticus | 1.17 | 0.28 - 4.8 | 0,832 | 0.74 | 0.11 - 4.86 | 0,751 |
| Tuberous sclerosis | 1.68 | 0.52 - 5.39 | 0,383 | 1.82 | 0.53 - 6.26 | 0,339 |
| LCM | **HR** | **95% CI** | **p** | **HR** | **95% CI** | **p** |
| Age epilepsy onset | 0.99 | 0.97 - 1.02 | 0,501 | 1.01 | 0.98 - 1.04 | 0,375 |
| Female | 1.01 | 0.81 - 1.25 | 0,937 | 1.36 | 0.82 - 2.26 | 0,24 |
| Nr previous ASMs | 1.07 | 1.02 - 1.12 | 0,004 | 1 | 0.81 - 1.25 | 0,981 |
| Brain infection | 0.59 | 0.31 - 1.1 | 0,099 | 1.06 | 1.01 - 1.12 | 0,022 |
| Brain tumor | 0.45 | 0.15 - 1.41 | 0,173 | 0.58 | 0.3 - 1.1 | 0,094 |
| Inborn errors of metabolism | 1.01 | 0.57 - 1.8 | 0,965 | 0.47 | 0.15 - 1.49 | 0,202 |
| Intellectual or developmental disorder | 1.31 | 1.05 - 1.64 | 0,018 | 1.05 | 0.59 - 1.89 | 0,867 |
| Neurological malformation | 1.01 | 0.76 - 1.34 | 0,95 | 1.28 | 1 - 1.63 | 0,055 |
| Status epilepticus | 0.77 | 0.43 - 1.38 | 0,383 | 0.92 | 0.68 - 1.23 | 0,56 |
| Tuberous sclerosis | 1.56 | 0.96 - 2.54 | 0,075 | 0.86 | 0.48 - 1.55 | 0,622 |
| PER | **HR** | **95% CI** | **p** | **HR** | **95% CI** | **p** |
| Age epilepsy onset | 1.02 | 0.97 - 1.06 | 0,492 | 1.02 | 0.97 - 1.08 | 0,402 |
| Female | 0.81 | 0.54 - 1.22 | 0,307 | 1.71 | 0.66 - 4.47 | 0,271 |
| Nr previous ASMs | 1 | 0.93 - 1.08 | 0,957 | 0.78 | 0.51 - 1.19 | 0,252 |
| Brain infection | 2.51 | 1.09 - 5.81 | 0,031 | 1.01 | 0.94 - 1.1 | 0,736 |
| Brain tumor | 2.55 | 0.62 - 10.47 | 0,193 | 2.83 | 1 - 8 | 0,05 |
| Inborn errors of metabolism | 0.49 | 0.16 - 1.56 | 0,231 | 2.71 | 0.61 - 12.04 | 0,19 |
| Intellectual or developmental disorder | 0.97 | 0.6 - 1.55 | 0,886 | 0.54 | 0.17 - 1.75 | 0,305 |
| Neurological malformation | 0.99 | 0.6 - 1.64 | 0,974 | 1.22 | 0.7 - 2.14 | 0,479 |
| Status epilepticus | 1.77 | 0.65 - 4.84 | 0,263 | 1.14 | 0.66 - 1.97 | 0,629 |
| Tuberous sclerosis | 1.57 | 0.64 - 3.88 | 0,324 | 0.96 | 0.29 - 3.19 | 0,95 |

eFigure 1. Number of Antiseizure medications tried before starting each of the newer ASMs (A) and age when starting the newer medications (B).

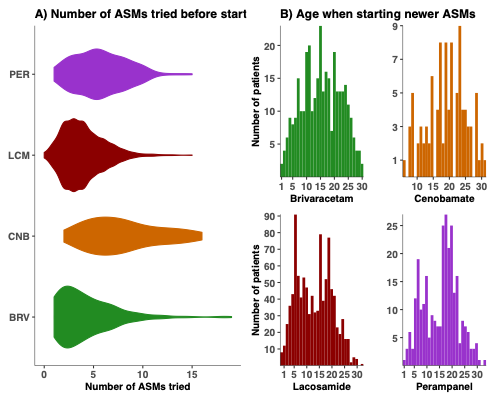
